# Vaccine Approvals and the Role of the FDA Vaccine Advisory Committee, 2000-2019

**DOI:** 10.1101/2021.07.19.21260761

**Authors:** Genevieve P. Kanter, Neel Vallurupalli, Yao Xu, Ravi Gupta

**Author notes:** Corresponding author:* Genevieve P. Kanter.

## Abstract

**Background:** The US Food and Drug Administration (FDA) plays a critical role in bolstering public confidence in vaccines and the vaccine review process. An important tool for enhancing transparency and public trust is the FDA’s Vaccine and Biological Related Products Advisory Committee (VRBPAC), a group of external experts that advises on scientific issues related to the licensure of vaccines.

**Objective:** To analyze key features of VRBPAC meetings convened over 20 years; estimate the probability of advisory committee review of newly approved vaccines, focusing on vaccines targeting emerging diseases; and examine the speed of and variance in approval times as a function of VRBPAC review.

**Methods:** Cross-sectional study of VRBPAC meetings convened and new vaccine licensure applications approved between January 1, 2000, and December 31, 2019. We analyzed the frequency of VRBPAC meetings and sessions; the percentage of newly licensed vaccines reviewed by VRBPAC; and the number of days between the submission of the licensure application and the date of FDA approval.

**Results:** Between 2000 and 2019, VRBPAC convened for a mean of 4.1 sessions per year. One-quarter of sessions was devoted to the review of specific vaccine products. During the same period, 44 new vaccine licensures were approved, 20% of which were for vaccines targeting emerging diseases. Almost half (48%) of successful new vaccine applications were reviewed by VRBPAC (n=21), a rate lower than for therapeutic applications. Among new applications targeting emerging diseases, 29% of non-influenza vaccines were reviewed by VRBPAC. There was no difference in the median time to approval as a function of VRBPAC review (364 days with VRBAC review vs. 365 days with no review, p=0.870).

**Conclusion:** The FDA has convened VRBPAC for reviews of about half of its vaccine products, less frequently for vaccines against non-influenza emerging diseases. There is considerable scope for the FDA to increase VRBPAC engagement in the vaccine review process.

## Introduction

Given the current fragile trust in COVID-19 vaccines,^1,2^ the US Food and Drug Administration (FDA) will play a critical role in bolstering public confidence in vaccines and the vaccine review process. One of the tools available to the FDA to strengthen public trust is its advisory committees, which are groups of external experts asked to advise on the approval of products reviewed by the agency. In the case of vaccines, the Vaccine and Biological Related Products Advisory Committee (VRBPAC) convenes regularly to advise on scientific issues related to the pre-market safety and efficacy of vaccines submitted for licensure, and review post-market safety. The independence of advisory committee experts and the public nature of VRBPAC meetings—which permit outside observers to view supporting clinical evidence, follow deliberations, and express their own views—are thought to bestow credibility on and further public trust in FDA decisions.^3-5^

The FDA, however, is not required to convene an advisory committee to review any vaccine candidate, nor is it required to follow VRBPAC recommendations.^6,7^ Consequently, there has been great discretion in the FDA’s historical use of VRBPAC. Now in the wake of the public trust challenges presented by COVID-19, the FDA appears poised to intensify its use of VRBPAC. Indeed, Dr. Peter Marks, the director of the FDA’s Center for Biologics Evaluation and Research has explicitly emphasized this trust role of the advisory committee and promised increased VRBPAC involvement: “ To ensure transparency regarding COVID-19 vaccines, the FDA intends to schedule meetings, as needed, of the VRBPAC… We recognize that being transparent about the data that we will evaluate in support of the safety and effectiveness of these vaccines and discussing this data with members of the VRBPAC in a public forum is critical to build trust and confidence in their use by the public.” ^8^

The increased engagement of VRBPAC for COVID vaccines and likely future vaccines in the face of increased vaccine hesitancy^9-11^ will affect vaccine researchers and developers, physicians, and the public. The increased probability of VRBPAC review will increase public scrutiny for certain types of vaccines and the evidence supporting them, and may extend review times. There has, however, been very little systematic examination of VRBPAC and its past involvement in vaccine licensures.

Previous research on FDA advisory committees has focused on financial conflicts of interest among committee members;^12-15^ concordance between advisory committee recommendations and final FDA decisions;^16,17^ and financial conflicts of interest among public speakers at committee meetings.^18-20^ There have also been case studies examining FDA approval of specific vaccines^21,22^ and summaries of FDA vaccine licensures.^23-25^ To date, however, there has been no comprehensive review of VRBPAC activities or analysis of VRBPAC involvement in vaccine approvals.

We analyze characteristics of VRBPAC meetings convened between 2000 and 2019 and estimate the probability of advisory committee review of vaccines approved during this 20-year period. We also compare the speed of and variance in approval times between vaccines that were reviewed by VRBPAC and those that were not. We examine these patterns for all new approved license applications and for the subset of vaccine applications targeting emerging diseases, which are likely to comprise an increasing share of new vaccines.^26^ Based on these analyses, we discuss implications for increased VRBPAC involvement.

## Methods

### Data

To obtain information on advisory committee activity, we extracted meeting characteristics from agenda and transcript documents publicly posted on the FDA website for all VRBPAC meetings convened between January 1, 2000, and December 31, 2019.^27^ From these documents, we collected the dates and session topics discussed at each meeting (there are typically multiple sessions during a single meeting), and identified whether the session was voting or non-voting, was open or closed to the public, and had any financial conflict of interest waivers issued. We grouped session topics into 8 categories: briefing related to research conducted by the Office of Vaccines Research and Review; review related to a specific vaccine product; seasonal flu strain selection; other flu update; safety or efficacy of vaccines under clinical development; safety or efficacy of currently marketed vaccines; vaccine production and manufacturing; and administrative (non-research) briefing. For product review meetings, we identified the specific product that was under review. We also identified meetings that discussed issues related to vaccines targeting emerging diseases as identified by the National Institute of Allergy and Infectious Diseases (NIAID).^28^ NIAID identifies diseases and pathogens that pose a risk to national security and public health because they present either a natural or deliberately released biological threat. These include, for example, anthrax, Ebola virus disease, pandemic influenza, and MERS-CoV.

To identify all vaccines licensed between 2000 and 2019, we first generated a list of vaccines based on the vaccine list posted on the “ Vaccines Licensed for Use in the United States” page of the FDA website.^29^ Because this page includes information only on currently licensed vaccines, we enriched our original list with vaccine lists from Pickering et al’s review of vaccines^24^ and the most recent edition of *Epidemiology and Prevention of Vaccine-Preventable Diseases*^30^ (“ Pink Book”) to identify vaccines that had been approved but withdrawn from the US market. For each of the vaccines listed on the FDA website, we used publicly posted approval documents to extract the Biological License Application (BLA) number and Submission Tracking Number of applications for new vaccine licenses; participation in any FDA expedited programs, including accelerated approval, fast track, breakthrough therapy, and priority review; application submission date(s); approval date; and indication. For the analysis of vaccines against emerging diseases, we also reviewed documents for BLA supplemental applications for expanded indications or expanded subtypes or variants.

For every product review meeting, we searched for BLA approvals of that product after the advisory committee meeting. If we found an approval for the product discussed, we reviewed the meeting transcript to confirm that VRBPAC had recommended the product for approval. If we were unable to find an approval for the product, we reviewed the meeting transcript to confirm that VRBPAC had not recommended the product for approval.

Because this study used publicly available data on meetings and vaccine approvals, it is not considered human subjects research.

### Outcomes

The main outcomes of interest were the frequency of VRBPAC meetings and meeting sessions, over time and by different characteristics (e.g., topics); the percentage of newly licensed vaccines that had been reviewed by the advisory committee; the number of days between the first submission of the licensure application and the date of FDA approval; and for those vaccines that had undergone VRBPAC review, the number of days between the VRBPAC meeting and the date of FDA approval.

### Statistical Methods

Because of the small sample sizes, classical hypothesis testing applied to most outcomes was uninformative (we note that the dataset is also a census of all vaccines approved and all meetings convened). For this reason, we primarily report summary statistics of meeting and vaccine characteristics. We calculated the probability of VRBPAC review as the percentage of approved vaccines that were reviewed by VRBPAC, both unconditionally and also stratifying by vaccine characteristics. In comparing the time to vaccine approval for vaccines that underwent VRBPAC review versus those that did not, we used a *t*-test to test for equality of means; an F-test to test for equality of variances; and a non-parametric χ^2^ test to test for equality of medians.

## Results

Between 2000 and 2019, the Vaccines and Related Blood Products Advisory Committee held 82 meetings, convening for 179 distinct sessions (Table 1). Most sessions (72%) were open to the public, and about half of the open sessions (46%) were voting sessions. Financial conflict of interest waivers were issued for 29% of open sessions.

**Table 1.** Characteristics of Vaccines and Related Blood Products Advisory Committee (VRBPAC) meetings and sessions, 2000-2019.

| Characteristic | Meetings<br>(n=82) | Sessions<br>(n=179) |
| --- | --- | --- |
| Years |  |  |
| 2000-2004 | 23 (28%) | 63 (35%) |
| 2005-2009 | 21 (26%) | 49 (27%) |
| 2010-2014 | 16 (20%) | 33 (18%) |
| 2015-2019 | 22 (27%) | 34 (19%) |
| Open or closed session |  |  |
| open |  | 128 (72%) |
| closed |  | 51 (28%) |
| Voting or non-voting session, open sessions only (n=128) |  |  |
| voting |  | 59 (46%) |
| non-voting |  | 69 (54%) |
| Financial conflicts of interest, open sessions only (n=128) |  |  |
| No waivers |  | 46 (36%) |
| Waivers issued |  | 37 (29%) |
| Not applicable |  | 45 (35%) |
| Topics (annual number of sessions: mean 4.1, median 4) |  |  |
| Research program briefing (mean 1.8, median 2) |  | 36 (28%) |
| Specific product (mean 1.6, median 1) |  | 32 (25%) |
| Seasonal flu strain selection (mean 1.5, median 1) |  | 30 (23%) |
| Safety/efficacy, vaccines under development (mean 0.8, median 1) |  | 15 (11%) |
| Vaccine production and manufacturing (mean 0.4, median 0) |  | 7 (5%) |
| Safety/efficacy, currently marketed vaccines (mean 0.2, median 0) |  | 4 (3%) |
| Administrative (non-research) briefing (mean 0.1, median 0) |  | 2 (2%) |
| Other flu update (mean 0.1, median 0) |  | 2 (2%) |

Over the 20-year period, VRBPAC convened for a mean of 4.1 open sessions (median 4 sessions) each year. The greatest number of sessions (n=36, 28%) was devoted to briefings on research programs conducted by the Office of Vaccines Research and Review. One-quarter of the sessions reviewed specific products, and 23% deliberated on the strains to be selected for the upcoming season’s influenza vaccines. VRBPAC met, on average, for 1.6 sessions per year to review specific vaccine products.

VRBPAC met for 12 sessions to discuss vaccines related to emerging diseases (S1 Table). These included 4 voting product review sessions (2 for H5N1 influenza, 1 for smallpox, 1 for dengue) and 6 non-product-specific sessions to discuss emerging threats (4 for pandemic influenza and 1 each for Ebola and chikungunya). There were also 2 sessions held to discuss the contemporaneous pandemic threat of H1N1 influenza in 2009, but neither session reviewed a specific product. The committee did not convene for any meetings related to Middle East Respiratory Syndrome (MERS), or Severe Acute Respiratory Syndrome (SARS), or Zika virus disease.

We identified 44 new vaccine licensure approvals between 2000 and 2019 (Table 2). The largest number of new licensures were for vaccines targeting NIAID emerging diseases, a heterogeneous group that included H5N1 influenza, cholera, dengue, Ebola, hepatitis A, Japanese encephalitis, and smallpox (n=9, 20%). Other common licensures were for vaccines against seasonal influenza (n=8, 18%), DTaP (n=6, 13%), and meningococcus (n=5, 11%). Fifty-nine percent of vaccine licensures were approved through the traditional pathway; 18% went through the accelerated pathway; 18% had fast track designation, and 27% received priority review.

**Table 2.**
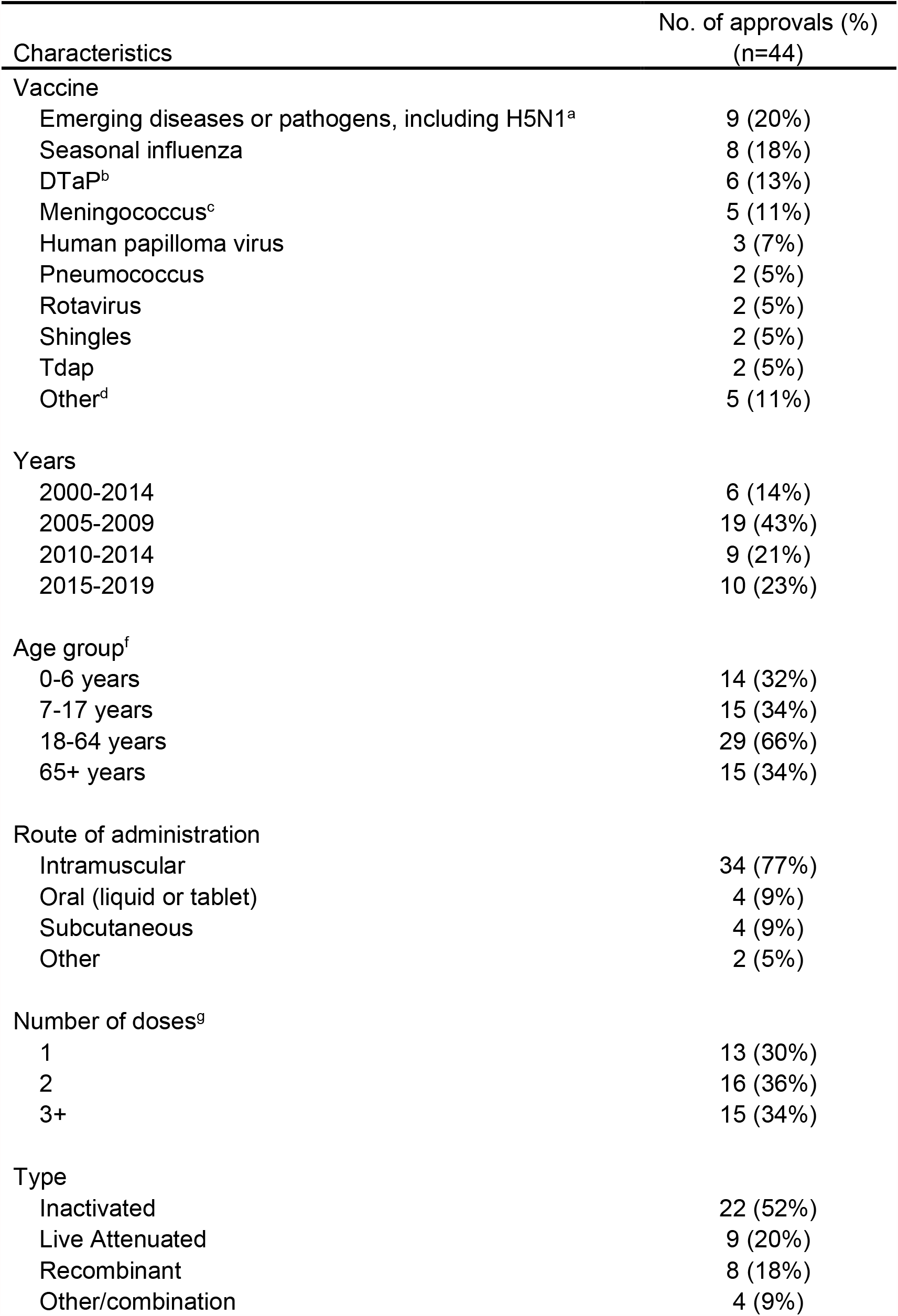
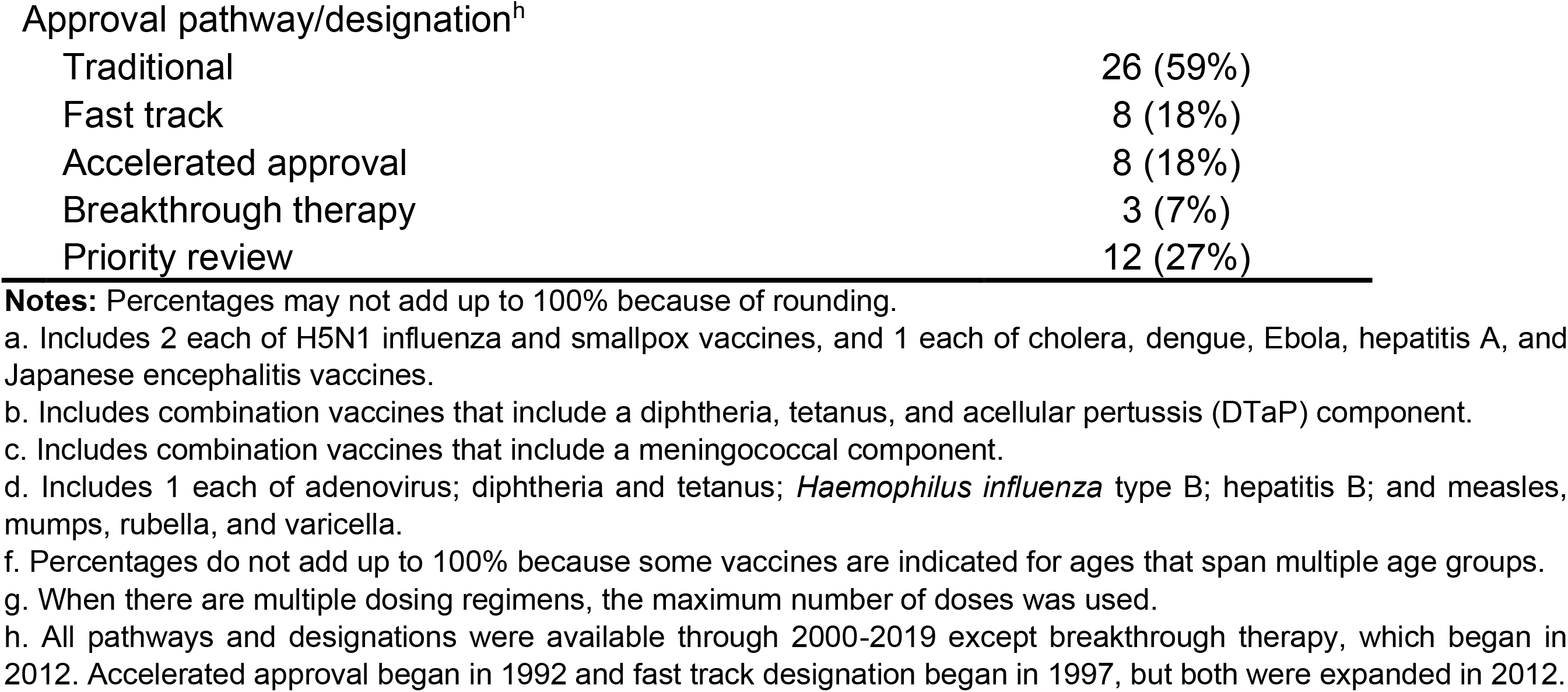
Characteristics of new vaccine approvals, 2000-2019.

| Characteristics | No. of approvals (%)<br>(n=44) |
| --- | --- |
| Vaccine |  |
| Emerging diseases or pathogens, including H5N1<sup>a</sup> | 9 (20%) |
| Seasonal influenza | 8 (18%) |
| DTaP<sup>b</sup> | 6 (13%) |
| Meningococcus<sup>c</sup> | 5 (11%) |
| Human papilloma virus | 3 (7%) |
| Pneumococcus | 2 (5%) |
| Rotavirus | 2 (5%) |
| Shingles | 2 (5%) |
| Tdap | 2 (5%) |
| Other<sup>d</sup> | 5 (11%) |
| Years |  |
| 2000-2014 | 6 (14%) |
| 2005-2009 | 19 (43%) |
| 2010-2014 | 9 (21%) |
| 2015-2019 | 10 (23%) |
| Age group<sup>f</sup> |  |
| 0-6 years | 14 (32%) |
| 7-17 years | 15 (34%) |
| 18-64 years | 29 (66%) |
| 65+ years | 15 (34%) |
| Route of administration |  |
| Intramuscular | 34 (77%) |
| Oral (liquid or tablet) | 4 (9%) |
| Subcutaneous | 4 (9%) |
| Other | 2 (5%) |
| Number of doses<sup>g</sup> |  |
| 1 | 13 (30%) |
| 2 | 16 (36%) |
| 3+ | 15 (34%) |
| Type |  |
| Inactivated | 22 (52%) |
| Live Attenuated | 9 (20%) |
| Recombinant | 8 (18%) |
| Other/combination | 4 (9%) |
| Approval pathway/designation<sup>h</sup> |  |
| Traditional | 26 (59%) |
| Fast track | 8 (18%) |
| Accelerated approval | 8 (18%) |
| Breakthrough therapy | 3 (7%) |
| Priority review | 12 (27%) |

**Notes:** Percentages may not add up to 100% because of rounding.
a. Includes 2 each of H5N1 influenza and smallpox vaccines, and 1 each of cholera, dengue, Ebola, hepatitis A, and Japanese encephalitis vaccines.
b. Includes combination vaccines that include a diphtheria, tetanus, and acellular pertussis (DTaP) component.
c. Includes combination vaccines that include a meningococcal component.
d. Includes 1 each of adenovirus; diphtheria and tetanus; *Haemophilus influenza* type B; hepatitis B; and measles, mumps, rubella, and varicella.
f. Percentages do not add up to 100% because some vaccines are indicated for ages that span multiple age groups.
g. When there are multiple dosing regimens, the maximum number of doses was used.
h. All pathways and designations were available through 2000-2019 except breakthrough therapy, which began in 2012. Accelerated approval began in 1992 and fast track designation began in 1997, but both were expanded in 2012.

Overall, almost half of successful new vaccine applications were reviewed by VRBPAC (n=21 out of 44, 48%). Fig 1 shows percentage of vaccines reviewed by VRBPAC, grouped by pathogen. There was no consistent pattern of VRBPAC review based on vaccine characteristics that were available prior to committee review. Fig 2 shows the probability of VRBPAC review stratified by various vaccine characteristics. The point estimates suggest little difference in the probability of review related to the type (e.g., live attenuated), indicated age group, or number of doses. However, approvals that went through some expedited pathways were less likely to have been reviewed by VRBPAC. While 54% of traditional approvals went to the advisory committee, 13% of accelerated approvals and 0% of breakthrough therapies did so.

**Figure 1.**
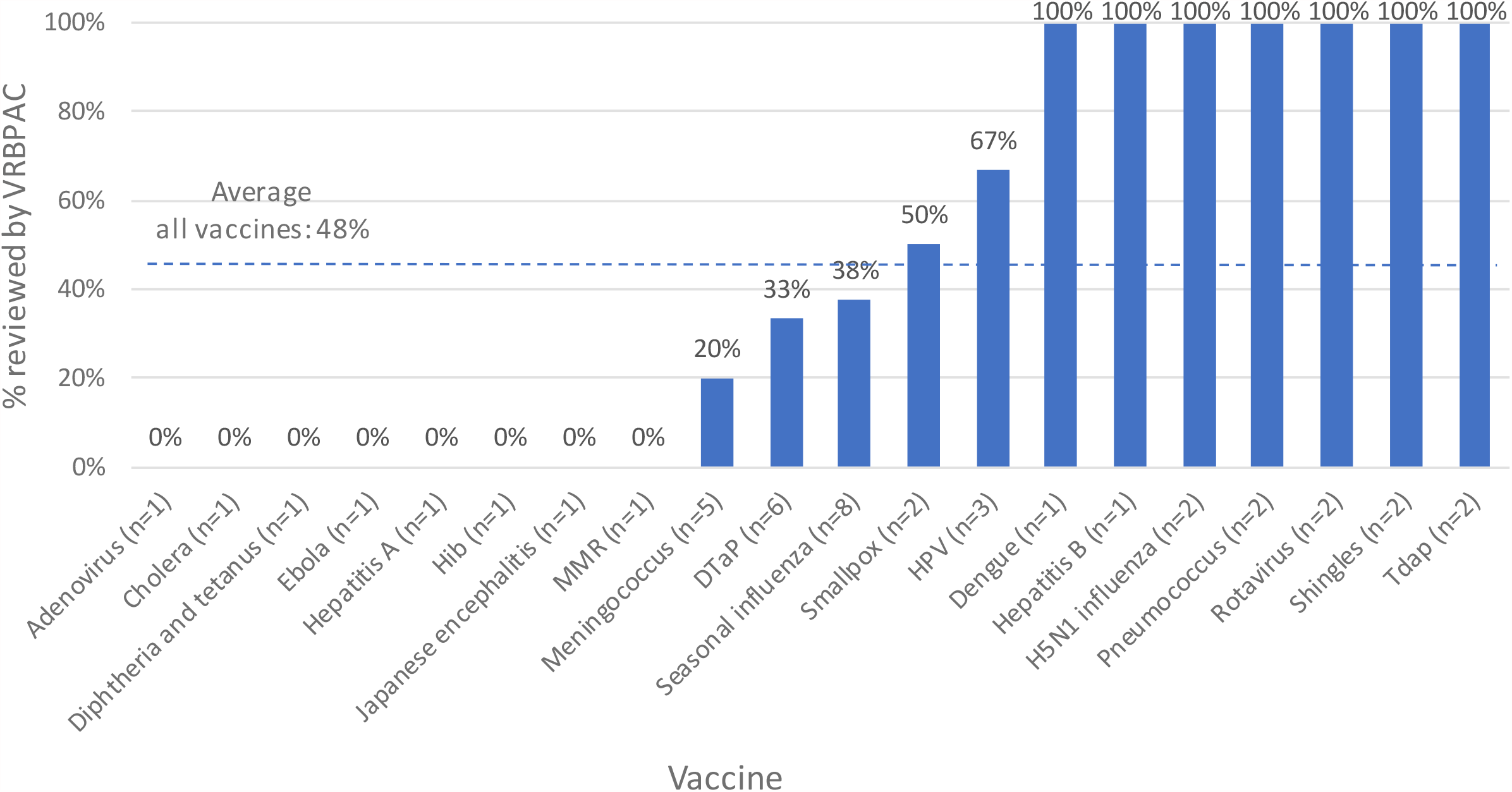
Percentage of newly licensed vaccines reviewed by VRBPAC, 2000-2019. **Notes:** The total number of vaccines of each type is reported in parentheses. The horizontal dashed line indicates the percentage of all newly licensed vaccines reviewed by VRBPAC (48%).

**Figure 2.**
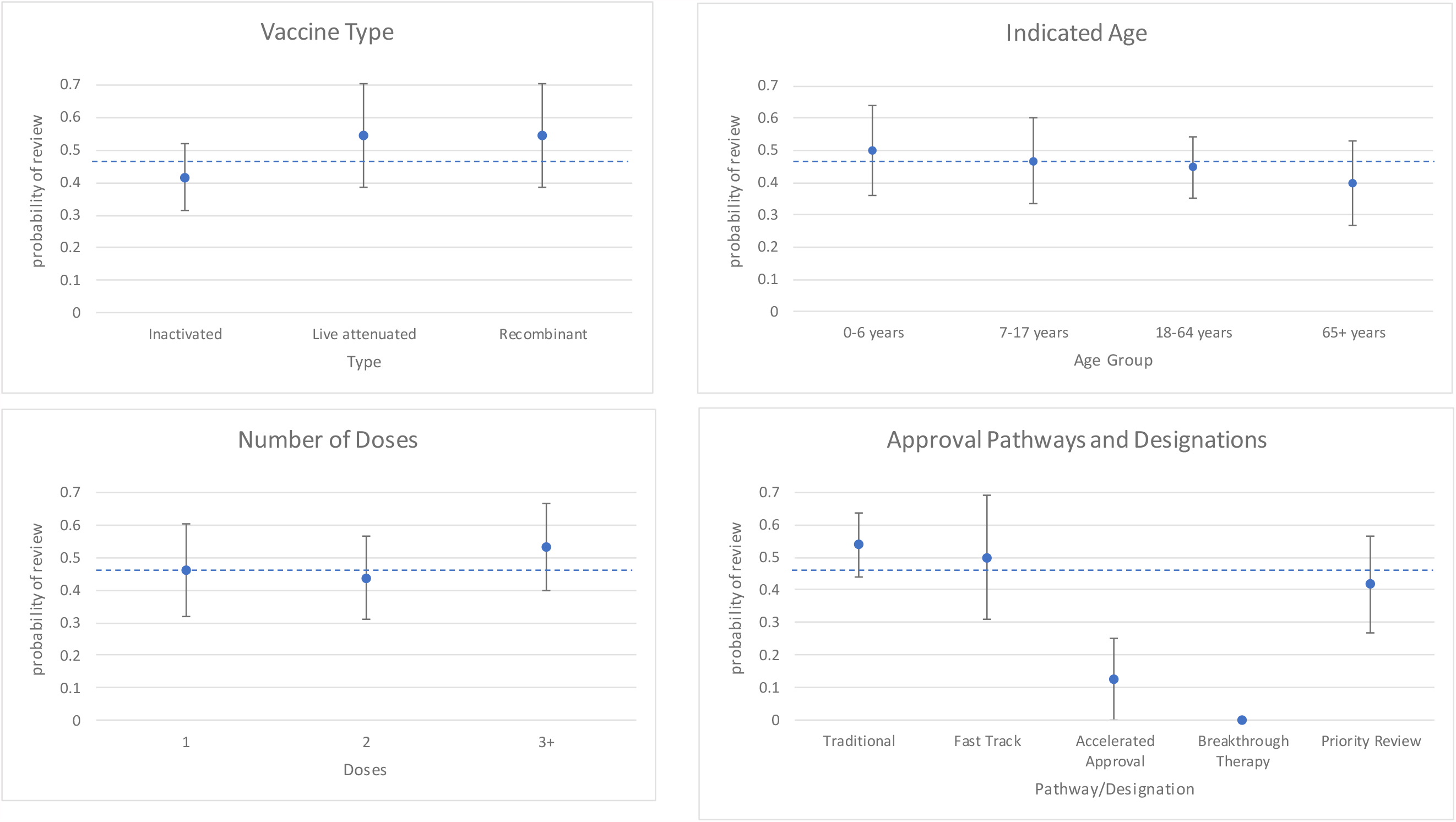
Probability of VRBPAC review of new vaccine approvals, by vaccine characteristics. **Notes:** Point estimates and standard errors are shown. The horizontal dashed line indicates the unconditional probability of VRBPAC review for all vaccines (48%). For vaccine type, the number of vaccines in each category is 23 (inactivated), 9 (live attenuated), and 7 (recombinant). For indicated age, the number of vaccines is 14 (0-6 years), 15 (7-17 years), 29 (18-64 years), and 15 (65+ years). Some vaccines are indicated for ages that span multiple groups. For number of doses, the number of vaccines is 13 (1 dose), 16 (2 doses), 15 (3+ doses). For approval pathways and designations, the number of vaccines is 26 (traditional), 8 (fast track), 8 (accelerated approval), 3 (breakthrough therapy), and 12 (priority review). Some vaccines have multiple designations.

Fourteen vaccines against emerging diseases were approved during this period, 9 as new licensures and 5 as supplements (Table 3). Of the new licensures, 4 (44%) were reviewed by VRBPAC. None of the supplements were reviewed by VRBPAC.

**Table 3.** FDA approval and VRBPAC review of vaccines for emerging diseases, 2000-2019.

| Vaccine | Original Sponsor | Approval Month/Year | New BLA or Supplement | VRBPAC review |
| --- | --- | --- | --- | --- |
| Hepatitis A | SmithKline Beecham | 5/2001 | New | no |
| Influenza H5N1 | Sanofi Pasteur | 4/2007 | New | yes |
| Smallpox | Acambis | 8/2007 | New | yes |
| Japanese encephalitis | Intercell | 3/2009 | New | no |
| Influenza H1N1 | Novartis | 9/2009 | Supplement | no |
| Influenza H1N1 | MedImmune | 9/2009 | Supplement | no |
| Influenza H1N1 | ID Biomedical | 11/2009 | Supplement | no |
| Japanese encephalitis | Intercell | 5/2013 | Supplement | no |
| Influenza H5N1 | ID Biomedical | 11/2013 | New | yes |
| Cholera | PaxVax | 6/2016 | New | no |
| Influenza H5N1 | ID Biomedical | 9/2016 | Supplement | no |
| Dengue | Sanofi Pasteur | 5/2019 | New | yes |
| Smallpox | Bavarian Nordic | 9/2019 | New | no |
| Ebola | Merck Sharp & Dohme | 12/2019 | New | no |

For applications that underwent VRBPAC review (Table 4), the mean time to approval was 659 days, compared to a mean time of 487 days for those applications that did not, but this difference was not statistically significant (172 days, 95% CI (−144,488), p=0.278). The variance in review times was larger for applications undergoing VRBPAC review than for those that did not (SD 610 days vs. 397 days); this difference was slightly above statistical significance (p=0.058). There was little difference in the median time to approval: 364 days for VRBPAC review vs. 365 days for no VRBPAC review (p=0.870). For applications reviewed by VRBPAC, the median number of days between VRBPAC review and FDA approval was 98 days, or about 3 months.

**Table 4.** Time to approval of new vaccine applications, 2000-2019.

|  | All<br>Applications<br>(n=43)<sup>a</sup> | VRBPAC<br>Review<br>(n=21)<sup>b</sup> | No VRBPAC<br>Review<br>(n=22)<sup>c</sup> | p-value |
| --- | --- | --- | --- | --- |
| Days between date of first submission and date of FDA approval |  |  |  |  |
| Mean | 571 | 659 | 487 | 0.278<sup>d</sup> |
| Standard deviation | 513 | 610 | 397 | 0.058<sup>e</sup> |
| Median | 365 | 364 | 365 | 0.870<sup>f</sup> |
| Interquartile range | 587 | 638 | 356 |  |
| Min | 98 | 182 | 98 |  |
| Max | 2,175 | 2,175 | 1,591 |  |
| Days between date of VRBPAC meeting and date of FDA approval<sup>g</sup> |  |  |  |  |
| Mean |  | 204 |  |  |
| Standard deviation |  | 270 |  |  |
| Median |  | 98 |  |  |
| Interquartile range |  | 131 |  |  |
| Min |  | 37 |  |  |
| Max |  | 1,154 |  |  |

**Notes:**
a. The dates of first submission for all but one vaccine were available from public documents.
b. 7 out of 21 applications that were reviewed by VRBPAC had some type of expedited designation.
c. 11 out of 22 applications that were not reviewed by VRBPAC had some type of expedited designation.
d. p-value from t-test of equality of means. 95% CI of difference in mean days: (-144,488).
e. p-value from F-test of equality of variances.
f. p-value from $\chi^2$ test of equality of medians.
g. If multiple VRBPAC meetings were convened to review a product, the date of the most recent VRBPAC meeting was used to calculate time to approval.

## Discussion

Over the past 20 years, the Vaccines and Related Products Advisory Committee has convened about 4 times a year, with most sessions devoted to routine topics such as selection of seasonal flu strains and briefings of research conducted by the FDA’s Office of Vaccines (the FDA conducts its own laboratory research on vaccines). On average, only 1.6 sessions per year were devoted to reviewing vaccine product applications and 0.8 sessions per year were spent discussing safety and efficacy issues related to vaccines under development. The frequency of vaccine product meetings is below the typical meeting frequencies observed for advisory committees for pharmaceuticals.^13,17^

During the same period, the FDA approved 44 new vaccine licensures. The frequency of vaccine licensure approvals is very low relative to approvals of new therapeutics. For comparison, 48 new pharmaceutical and biological therapeutics were approved in 2019 alone.^31^ These low rates of vaccine licensure are consistent with the relatively meager commercial investment in and financial returns from vaccines compared to therapeutics.^32^

The probability of a newly licensed vaccine having undergone VRBPAC review was 48% for 2000-2019. This probability of advisory committee review is somewhat lower than probabilities observed for advisory committee reviews of pharmaceuticals, which ranged from 67% to 95% in 1985-1999 and 50%-77% in 2000-2005.^33^

Of the 14 vaccine applications approved against emerging diseases, only 3 went to VRBPAC for review. Six of these vaccines were for pandemic (H1N1 and H5N1) strains of influenza, most of which were approved as supplements to existing flu vaccines. Supplements, even for expanded indications and strains, are generally considered to be lower risk and are far less likely to be sent for advisory committee review. Two H5N1 applications were new submissions, and both of these underwent VRBPAC review. However, only 29% of new applications for non-influenza emerging diseases were reviewed by VRBPAC.

Between 2000 and 2019, VRBPAC convened 8 sessions to discuss general (non-product-specific) issues about vaccines against emerging diseases. All but 2 were related to pandemic influenza. Notably, VRBPAC did not convene any meetings related to the MERS, SARS, or Zika pandemics. The FDA thus appears to have engaged more extensively with VRBPAC for vaccines related to pandemic influenza than for any other pandemic pathogens.

Historically, median times have not differed between applications that underwent VRBPAC review compared to those that did not; both routes took about a year. This is surprising because vaccines that would have been chosen for VRBPAC review would be expected to present more complicated safety or efficacy evidence—hence the need for external advice—so deliberations on the balance of benefits and harms would have taken longer. On the other hand, products are often be brought to the advisory committee for non-scientific reasons—such as a desire to divert any backlash from licensing a politically contentious vaccine to an external body^34^ or to demonstrate transparency. These latter rationales would not necessarily extend review times.

There was, however, somewhat greater variance in review times for applications reviewed by VRBPAC compared to those that were not. This suggests a greater risk of approval delay if the sponsor’s application is directed to the advisory committee. These delays may well involve requests for additional studies and data, and multiple VRBPAC reviews.

Given (1) the historically infrequent use of VRBPAC in product review, especially relative to advisory committees for therapeutics; (2) increased concerns about readiness for emerging diseases in light of COVID-19; (3) a disproportionate amount of VRBPAC meeting time spent on routine matters that could be redirected towards discussion of pre-market vaccine development; and (4) increasing vaccine hesitancy and fragility of public trust, the FDA has the scope and is likely to see a need for increasing its engagement with VRBPAC. This need not result in significantly longer review times for the typical applicant. Sponsors may, however, pre-emptively collect additional data or make other efforts to prevent embarrassing delays that might come from public scrutiny. While these activities would not extend the FDA review period, they would extend the clinical development period.

The COVID-19 pandemic has challenged many aspects of FDA operations. Despite facing unprecedented political and scientific hurdles, the agency has responded with greater transparency in its public review of SARS-CoV-2 vaccines than its own history would have predicted. In convening VRBPAC meetings to review the evidence for the emergency use authorization (EUA) of SARS-CoV-2 vaccines, the FDA has shown a willingness to deploy VRBPAC review to bolster trust in the review process and in the safety and efficacy of any authorized vaccines. Prior to COVID-19, there had only been 1 EUA of a vaccine—the anthrax vaccine in 2005—and FDA issued that authorization without seeking advice from VRBPAC.

The FDA has publicly affirmed its commitment to a transparent process of COVID-19 vaccine review.^8,34^ This decision could set an important precedent for vaccine review that may only slightly delay review processes but will be critical for enhancing transparency and build trust in vaccines.

## Data Availability

All data used are publicly available. FDA advisory committee meeting materials are available at: https://www.fda.gov/advisory-committees/committees-and-meeting-materials. Vaccines licensed for use in the US are available at: https://www.fda.gov/vaccines-blood-biologics/vaccines/vaccines-licensed-use-united-states.

## Acknowledgments

We thank Dr. Joshua Sharfstein, Dr. Jason Schwartz, and Dr. Joseph Ross for their expertise and valuable advice on this manuscript. They received no financial compensation for their contributions. We thank Laquesha Sanders, John Barnes, Kathryn Cowie, Alice Rossmann, Alisa Bhakta, and Olivia Darko for their assistance with data collection; they received wage compensation for their services.

## SUPPORTING INFORMATION

**S1 Table.** VRBPAC sessions related to emerging diseases, 2000-2019.

| Date | Topic | Product-specific |
| --- | --- | --- |
| 2/27/2007 | safety and efficacy of an H5N1 inactivated influenza vaccine | yes |
| 2/27/2007 | clinical development of influenza vaccines for pre-pandemic uses | no |
| 5/17/2007 | safety and immunogenicity of live vaccinia virus smallpox vaccine | yes |
| 2/21/2008 | clinical development of influenza vaccines for pandemic and pre-pandemic uses | no |
| 2/19/2009 | clinical studies with pandemic influenza vaccines in pediatric population in absence of pandemic (H5N1) | no |
| 7/23/2009 | discussion of clinical trials to support use of vaccines against the 2009 H1N1 influenza virus | no |
| 11/18/2009 | update on FDA's influenza A (H1N1) 2009 monovalent vaccine activities; postmarket surveillance | no |
| 2/29/2012 | licensure pathways for pandemic influenza vaccines | no |
| 11/14/2012 | safety and immunogenicity of an influenza A (H5N1) virus monovalent vaccine | yes |
| 5/12/2015 | development and licensure of Ebola vaccines | no |
| 3/7/2019 | safety and efficacy of dengue tetravalent vaccine | yes |
| 11/8/2019 | development of chikungunya vaccines | no |

## Notes

### Competing Interest Statement

The authors have declared no competing interest.

### Funding Statement

This research was not suppported by external funding.

